# RT-LAMP-CRISPR-Cas13a technology as a promising diagnostic tool for the SARS-CoV-2 virus

**DOI:** 10.1101/2022.06.29.22277060

**Authors:** Concha Ortiz-Cartagena, Laura Fernández-García, Lucia Blasco, Olga Pacios, Inés Bleriot, María López, Rafael Cantón, María Tomás

## Abstract

At the end of 2019, the new coronavirus, SARS-CoV-2, began a pandemic that persists to date and which has caused more than 6.2 million deaths. In the last couple of years, researchers have made great efforts to develop a diagnostic technique that maintains high levels of sensitivity and specificity, since an accurate and early diagnosis is required to minimize the prevalence of SARS-CoV-2 infection. In this context, CRISPR-Cas systems are proposed as promising tools for development in diagnostic techniques due to their high specificity, highlighting that Cas13 endonuclease discriminates single nucleotide changes and displays a collateral activity against single stranded RNA molecules. With the aim of improve the sensitivity of the diagnosis, this technology is usually combined with isothermal pre-amplification reactions (SHERLOCK, DETECTR). Basing on this, we have developed an RT-LAMP-CRISPR-Cas13a for SARS-CoV-2 virus detection in nasopharyngeal samples without using RNA extraction kit that exhibited 100 % specificity and 83 % sensitivity, as well as a positive predictive value of 100 % and a negative predictive value of 100%, 81%, 79.1% and 66.7 % in <20 Ct, 20-30 Ct, >30 Ct and total Ct values, respectively.

**Importance:** During Covid19 crisis has driven the development innovative molecular diagnose including the CRISPR-Cas technology. This work we have performed a protocol working with RNA-extraction kit free samples, places RT-LAMP-CRISPR-Cas13a technology at the top of rapid and specific diagnostic methods for COVID19 due to the high levels of specificity (100%), sensitivity (83%), PPV (100%) and NPV (81% in high loads viral) obtained in clinical samples.

## Introduction

Since their emergence at the beginning of the 21^st^ century, coronaviruses have been recognized as a health concern because of their ability to cause severe respiratory infections in humans. At the end of 2019, a new coronavirus appeared, SARS-CoV-2, producing a novel illness, COVID-19, and showing two remarkable characteristics. On the one hand, the virus causes the development of an unusual viral pneumonia and, on the other hand, it is highly transmissible, and thus spreads rapidly [1-3]. This led to the SARS-CoV-2 pandemic that persists to date and which has caused more than 6.2 million deaths (WHO COVID-19 Dashboard https://covid19.who.int/).

Fortunately, vaccination campaigns have decreased the incidence of COVID-19 [4]. However, specialists claim that this virus is likely to coexist with us for a long time as the price of vaccines and the stability-necessary cold chain make it difficult for the vaccine to reach the most remote places in the world, as SARS-CoV-2 does. Together with the fact that no efficient therapy has been developed for COVID-19, this indicates that accurate and early diagnosis in point of care (POC) testing is required to minimize the prevalence of SARS-CoV-2 infection [1-3].

In the last couple of years, researchers have made great efforts to develop a diagnostic technique that maintains high levels of sensitivity and specificity, without the need for expensive equipment or highly trained personnel for its implementation. Such a diagnostic technique would allow the detection of SARS-CoV-2 infection in health centres, as well as at home or in the field, which would accelerate the identification of infected patients, enabling prompt treatment and halting the spread of SARS-CoV-2 worldwide [5].

The use of nucleic-acid as biomarkers has become the diagnostic gold standard, because of the species specificity of the technique and because DNA and RNA can be amplified [6].

Although detection of nucleic acids has been linked to the polymerase chain reaction (PCR) and quantitative PCR (qPCR), these amplification methods increase the associated costs and must be carried out by specialized personnel. Consequently, isothermal amplification reactions are becoming more important in the diagnosis of COVID-19 [5]. Although different methods of isothermal amplification are available, recombinase polymerase amplification (RPA) and loop-mediated isothermal amplification (LAMP) reactions are the most commonly used in research. The LAMP-based technique has displayed greater specificity than RPA [5, 7]. LAMP had previously been used to detect several microorganisms, and the aforementioned mentioned advantages led to its optimization for COVID-19 diagnosis, as it has been applied in association with other techniques which increase the diagnostic specificity, such as clustered regularly interspaced short palindromic repeats (CRISPR)-associated protein (CRISPR-Cas) systems [5, 8-10].

Naturally, CRIPSR-Cas systems provide adaptive immunity for bacteria and archaea, as they collect genomic fragments (spacers) from foreign elements (bacteriophages, plasmids and other mobile genetic elements), which are expressed in an RNA molecule form (crRNA) that guides an endonuclease protein (Cas) to the pathogen, for final degradation of its nucleic-acid material [11, 12].

Since their discovery, CRISPR-Cas systems have revolutionized the field of molecular biology. Initially they were presented as highly specific tools for genome editing. However, they are also applicable for the diagnosis and treatment of infectious diseases and are now considered key for development in these areas [11, 12].

Class 2 CRISPR-Cas systems have a simpler effector structure, which makes them more attractive for use in genome editing, diagnosis and treatment. In this class, Cas12 and Cas13 proteins display non-specific endonuclease activity when activated (collateral activity) against single stranded DNA (ssDNA) and RNA (ssRNA) respectively. This feature could be applied in clinical diagnosis, taking advantage of the reporter molecule target of this activity (collateral-based detection), which acts by amplifying the detection signal. Therefore, Cas12 and Cas13 are proposed as the most promising tools for use in diagnostic techniques, with the latter being particularly important in terms of specificity as it has the ability to discriminate single nucleotide changes [11].

Researchers recently developed two novel assays for detecting SARS-Cov-2 based on CRISPR-Cas technology: DETECTR and SHERLOCK. The DETECTR technique uses reverse transcription-LAMP (RT-LAMP) for amplification and Cas12 as an endonuclease, while SHERLOCK uses RT-RPA for amplification and Cas13 [13, 14]. On the basis of these works, in this study we describe the development and optimization of a LAMP-CRISPR-Cas13a technique for the diagnosis of SARS-CoV-2 infection in clinical samples in a process that does not require RNA-extraction or purification. With this technique, high levels of sensitivity and specificity, comparable to those associated with qPCR, were obtained.

## Material and methods

### Study of the state of the art

A study of the state of the art was conducted with the aim of comparing the use of different novel diagnostic techniques. First, we conducted a search in PubMed with “qPCR diagnosis COVID19” as keywords and compared the output with the number of publications on RT-LAMP and RT-LAMP-CRISPR strategies for COVID-19 diagnosis [15]. Finally, we collected data on the different sensitivity, specificity, positive predictive value (PPV) and negative predictive value (NPV) from 10 papers related to RT-LAMP and 10 papers on the RT-LAMP-CRISPR-Cas COVID19 diagnostic technique [16-35]. We used the results to calculate the parameters needed for the comparison.

### In silico analysis and design of RT-LAMP primers, crRNAs and RNA reporters

The nucleocapside gene (ID: ON394272.1) of the SARS-CoV-2 virus was selected for study. The target sequence was analyzed in silico with the aim of designing specific primers for amplification of a genetic region without any previously described mutation (N gene region 12-213 pb named as N2 gene) [36]. Three pairs of LAMP primers were designed using the PrimerExplorer V5 software (F3-B3, FIP-BIP, Floop-Bloop) to amplify the SARS-CoV-2 N2 gene. The FIP LAMP primer contained the T7 polymerase promotor in its sequences for the subsequent transcription step (Table 1).

**Table 1.**
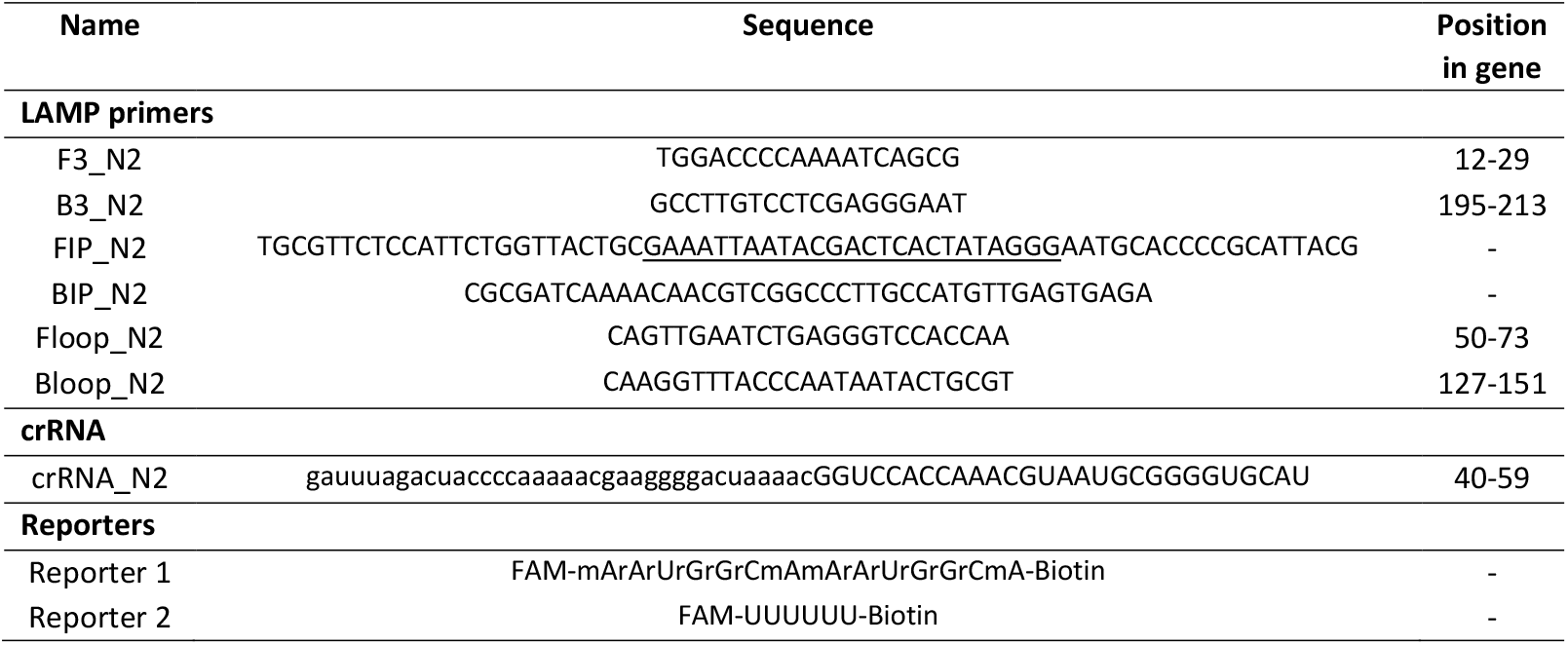
Sequences of primers, crRNAs and reporters. Underlined letters indicate overhang T7 promotor sequence and lower-case letters indicate scaffold sequence. All primers were supplied by IDT and reporters were supplied by GenScript.

Two different RNA reporters (reporter 1 and 2) were used to reveal the results in order to select the one with the best signal. Both contained a single isomer derivative of fluorescein modification (FAM) at the 5’
sextreme and a biotin molecule at the 3’extreme (Table 1).

### Clinical samples

Clinical samples were supplied by the Microbiology Service of the Teresa Herrera Materno Infantil Hospital (A Coruña, Spain). The samples (n= 133) were obtained from nasopharyngeal swabs for SARS-CoV-2 detection (Table 2).

**Table 2.**
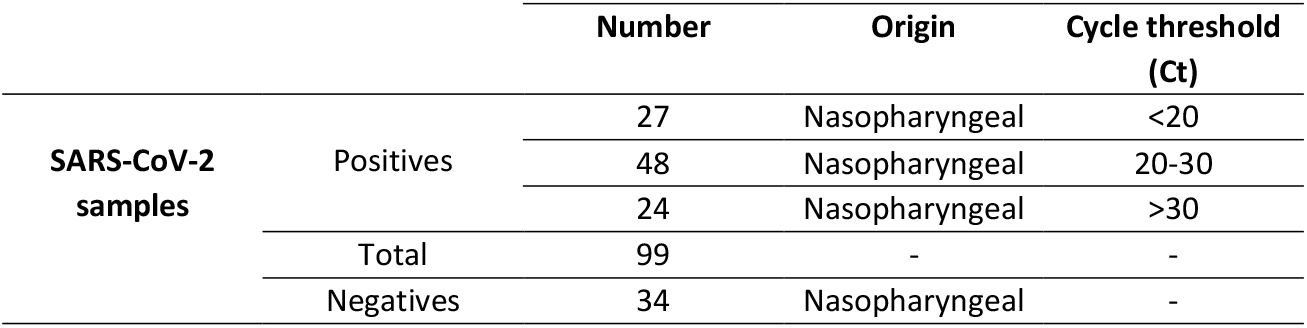
Positive and negative samples for SARS-CoV-2.

### Sample processing

For sample processing, a proteinase K-heat inactivation protocol (PK-HID) was applied to samples from swabs stored in viral transport medium (Gibco^®^) [37]. Thus, 95 μl aliquots of samples were treated for 15 minutes at 55°C with 5 μl of proteinase K (10 mg/ml, stock) prepared at 1 mg/ml in 100 μl of final volume and heat-inactivated at 98°C for 5 minutes. Finally, extracted RNA samples were stored at -80°C.

### RT-LAMP reaction

Amplification by the RT-LAMP (WarmStart^®^ LAMP Kit (DNA & RNA), NEB) reaction was performed following the manufacturer’s protocol. Briefly, RNA samples (5 μl) were added to a reaction mix containing 12.5 μl of WarmStart LAMP 2X Master Mix and 2.5 μl of Primer Mix 10X (FIP/BIP 16 μM, F3/B3 2 μM, LOOPF/LOOPB 4 μM, stock) adjusted to a final volume of 25 μl with dH_2_O. The reactions were incubated at 65°C for 1 hour.

### Collateral-based detection

Each Cas13a-based detection reaction was incubated at 37°C for 30 min with the following reaction components: 2 μl of cleavage buffer 10X (200 mM HEPES, 90 mM magnesium chloride, 600 mM sodium chloride), 0.5 μl of dNTPs (HiScribe™ T7 Quick High Yield RNA Synthesis Kit), 0.5 μl of T7 polymerase (HiScribe™ T7 Quick High Yield RNA Synthesis Kit), 20 U of RNase murine inhibitor (NEB), 0.15 μl Cas13a endonuclease (25 nM, MCLAB), 0.5 μl crRNA (50 nM, IDT), 2 μl of reporter (1000 nM, IDT) and 5 μl of cDNA sample, adjusted to a final volume of 20 μl with dH_2_O.

Different concentrations of Cas13a and crRNA (200, 100, 50 nM) were tested, and two different enzyme:guide molar ratios were used (1:1 and 2:1).

### Hybridetect lateral flow

Results were revealed by the HybriDetect lateral flow assay as described by the manufacturer (Milenia Biotec), with some modifications. Briefly, 20 μl of collateral-based detection product was mixed with 80 μl of assay buffer in a 96-well plate. Immediately, the gold-extreme of the trip was submerged in the mix and held for 2-3 minutes.

Following the manufacturer’s instructions, reactive strips required calibration before application for management of an optimal RNA reporter concentration and, as mentioned, reporters 1 and 2 were tested. Results obtained with two different assay buffers were also compared: the kit assay buffer and the same supplemented with 5% polyethylenglycol (PEG).

The results obtained, i.e. true positive (TP), false positive (FP), false negative (FN) and true negative (TN), were used to calculate the following parameters: sensitivity (TP/TP+FN), specificity (TN/TN+FP), positive predictive value (TP/TP+FP) and negative predictive value (TN/TN+FN).

### Limit of detection

For estimating the number of initial SARS-CoV-2 viral particles that the CRISPR-Cas13a technology was able to detect, we serially diluted (1:10) the RNA extracted using hospital equipment from two clinical samples with Ct 20 and Ct 25. Finally, 5 μL aliquots of each dilution were used for calculation of the limit of detection (LOD). Here, we applied an estimated correlation between Ct value and the viral load.

### Statistical analysis

Statistical analysis was conducted using the GraphPad Prism9 program to construct a receiver operating characteristic (ROC) curve with a confidence interval of 95% (Wilson/Brown method) and to construct a scatter plot of two groups (false negative and true positive samples) against the Ct of each sample.

## Results

### Analysis of the state of the art

We obtained an output of more than 7000 articles as a result of “qPCR diagnosis COVID19” search, of almost 4000 in 2021 alone. This result was compared with findings of Bhatt A. et al. [15] concerning papers related to RT-LAMP and CRISPR for SARS-CoV-2 diagnosis. Of these, we analyzed 10 articles on the RT-LAMP technique and 10 articles related to RT-LAMP-CRISPR-Cas technology, finding that only 1 applied the endonuclease Cas13 for SARS-CoV-2 diagnosis always on samples treated with an RNA extraction kit [35] (Table 3).

**Table 3.**
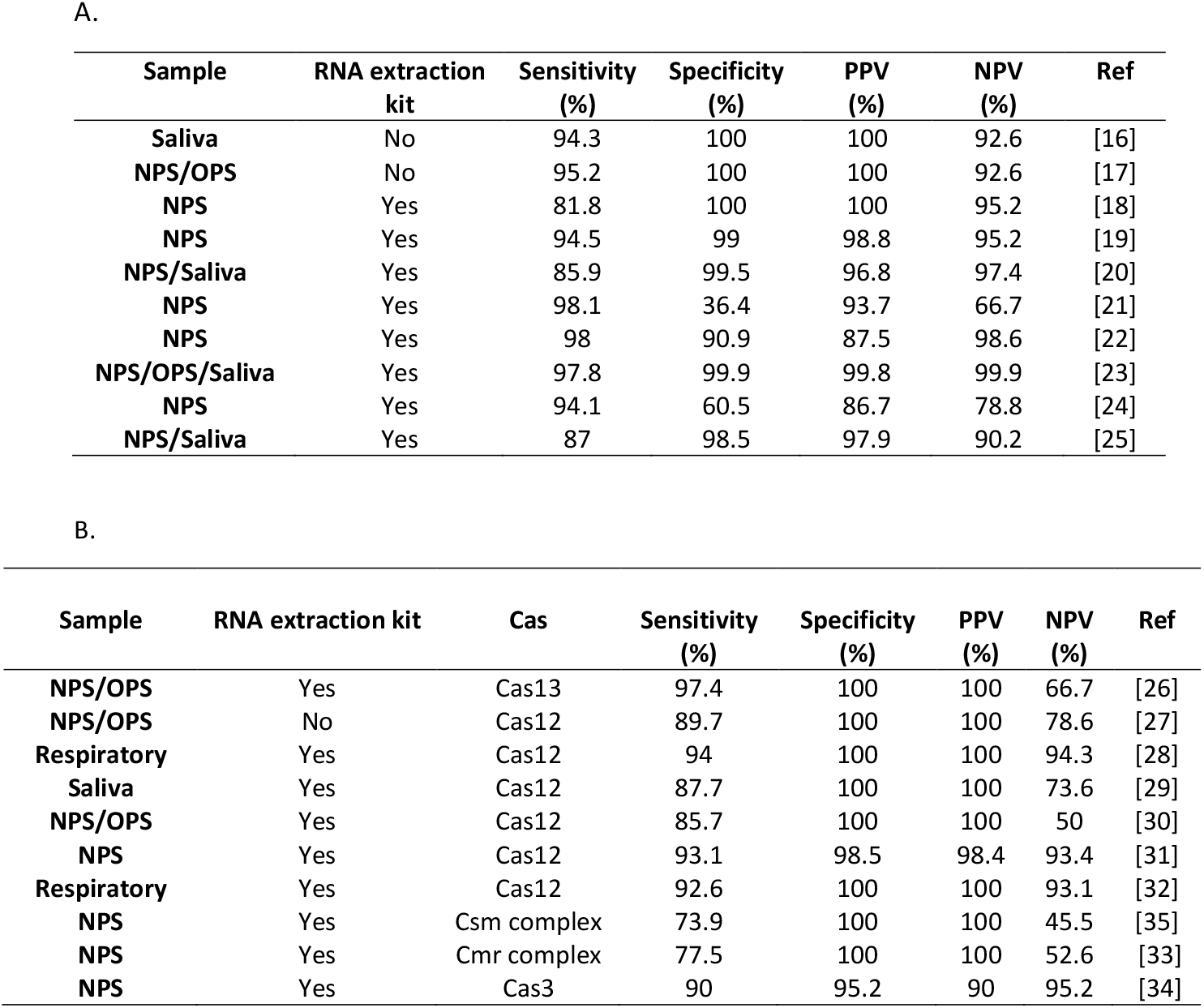
A. Table with percentages of sensitivity, specificity, PPV and NPV parameters collected from 10 articles which applies RT-LAMP technology to detect SARS-CoV-2 infection; B. Table with percentages of sensitivity, specificity, PPV and NPV parameters collected form ten articles which applies RT-LAMP-CRISPR technology to detect SARS-CoV-2 infection. Nasopharyngeal swab (**NPS**) and Oropharyngeal Swab (**OPS**).

| Sample | RNA extraction kit | Sensitivity (%) | Specificity (%) | PPV (%) | NPV (%) | Ref |
| --- | --- | --- | --- | --- | --- | --- |
| Saliva | No | 94.3 | 100 | 100 | 92.6 | [16] |
| NPS/OPS | No | 95.2 | 100 | 100 | 92.6 | [17] |
| NPS | Yes | 81.8 | 100 | 100 | 95.2 | [18] |
| NPS | Yes | 94.5 | 99 | 98.8 | 95.2 | [19] |
| NPS/Saliva | Yes | 85.9 | 99.5 | 96.8 | 97.4 | [20] |
| NPS | Yes | 98.1 | 36.4 | 93.7 | 66.7 | [21] |
| NPS | Yes | 98 | 90.9 | 87.5 | 98.6 | [22] |
| NPS/OPS/Saliva | Yes | 97.8 | 99.9 | 99.8 | 99.9 | [23] |
| NPS | Yes | 94.1 | 60.5 | 86.7 | 78.8 | [24] |
| NPS/Saliva | Yes | 87 | 98.5 | 97.9 | 90.2 | [25] |

| Sample | RNA extraction kit | Cas | Sensitivity (%) | Specificity (%) | PPV (%) | NPV (%) | Ref |
| --- | --- | --- | --- | --- | --- | --- | --- |
| NPS/OPS | Yes | Cas13 | 97.4 | 100 | 100 | 66.7 | [26] |
| NPS/OPS | No | Cas12 | 89.7 | 100 | 100 | 78.6 | [27] |
| Respiratory | Yes | Cas12 | 94 | 100 | 100 | 94.3 | [28] |
| Saliva | Yes | Cas12 | 87.7 | 100 | 100 | 73.6 | [29] |
| NPS/OPS | Yes | Cas12 | 85.7 | 100 | 100 | 50 | [30] |
| NPS | Yes | Cas12 | 93.1 | 98.5 | 98.4 | 93.4 | [31] |
| Respiratory | Yes | Cas12 | 92.6 | 100 | 100 | 93.1 | [32] |
| NPS | Yes | Csm complex | 73.9 | 100 | 100 | 45.5 | [35] |
| NPS | Yes | Cmr complex | 77.5 | 100 | 100 | 52.6 | [33] |
| NPS | Yes | Cas3 | 90 | 95.2 | 90 | 95.2 | [34] |

Data collected from RT-LAMP articles were used to determine the ranges of values of the parameters considered: sensitivity, 81 % -98 %; specificity, 36 % -100 %; PPV, 86 % -100 %; and NPV, 78 % -99 % (Table 3A). The results showed that major efforts have been made to detect SARS-CoV-2 virus in RNA purified samples (8/10), although RNA extraction-free research has also yielded potentially useful results (sensitivity >94 %, specificity and PPV 100 %, NPV >92 %). However, the highest levels of sensitivity and specificity were obtained in the projects involving extracted viral RNA (Table 3A).

Most reviewed papers (8/10) related to RT-LAMP-CRISPR-Cas technology used samples treated with extraction kits. Moreover, only 1 study applied the Cas13 enzyme as an effector protein and used RNA extracted by kit. In this case, the values for calculated data were 73 %-97 % for sensitivity, 95 %-100 % for specificity, 90 %-100 % for PPV and 50 %-95 % for NPV (Table 3B).

### SARS-CoV-2 detection

The best results for collateral-based detection reaction were achieved with 50 nM Cas13a enzyme and a molar ratio for Cas13a:crRNA of 2:1. On the other hand, HybriDetection lateral flow showed the higher sensitivity when reporter 2 was used at a final concentration of 1000 nM and the assay buffer was supplemented with 5 % PEG.

The LOD determination of the CRISPR-Cas13a based technology revealed that this technique detects as few as 1-10 SARS-CoV-2 particles (Figure 2). After PK-HID treatment, the LAMP-CRISPR-Cas13a technique was able to correctly detect samples with Ct<20 as positive. From samples with Ct 20-30 and Ct>30, the technique identified coronavirus as present in 83.3 % and 62.5 % of the samples, respectively (Figure 3C). Finally, the CRISPR-Cas13a technology did not detect SARS-CoV-2 infection in negative samples (Figure 3A). Based on the results obtained (Figure 3B), we estimated that the LAMP-CRISPR-Cas13a method for COVID-19 detection exhibits 100 % specificity and 83 % sensitivity, as well as a positive predictive value (PPV) of 100 % and a negative predictive value (NPV) of 100%, 81%, 79.1% and 66.7 % in <20 Ct, 20-30 Ct, >30 Ct and total Ct values, respectively (Figure 3C). The statistical analysis yielded a ROC curve with an area under the curve (AUC) of 0.84 with a 95 % CI of 0.73-0.93 (Figure 4A); in addition, examination of the scatter plot revealed that diagnostic results could be confused in nasopharyngeal samples with Ct>30 (Figure 4B).

**Figure 1.**
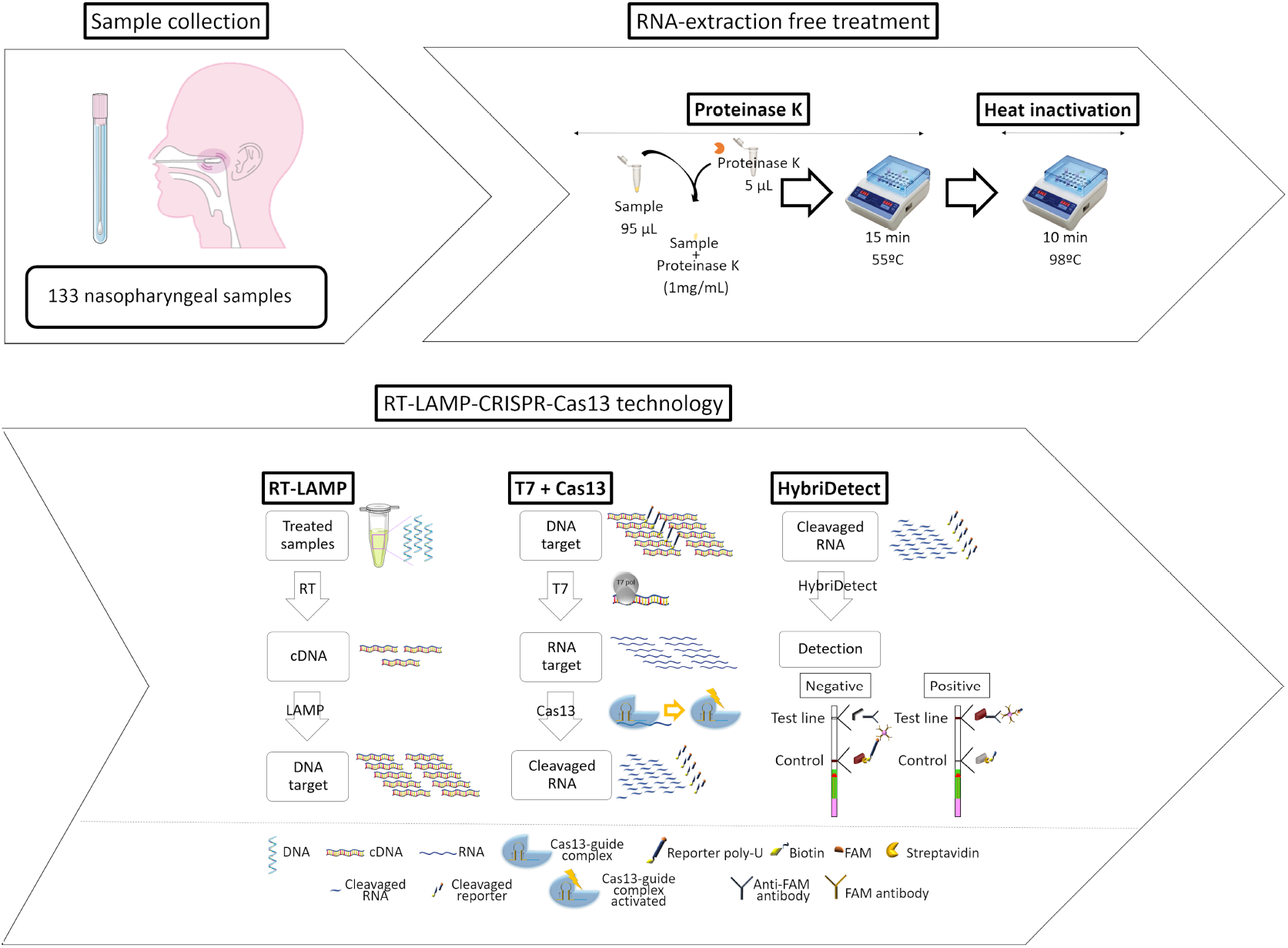
Workflow of the novel developed and optimized protocol for infectious diseases diagnosis based on CRISPR-Cas13a technology.

**Figure 2.**
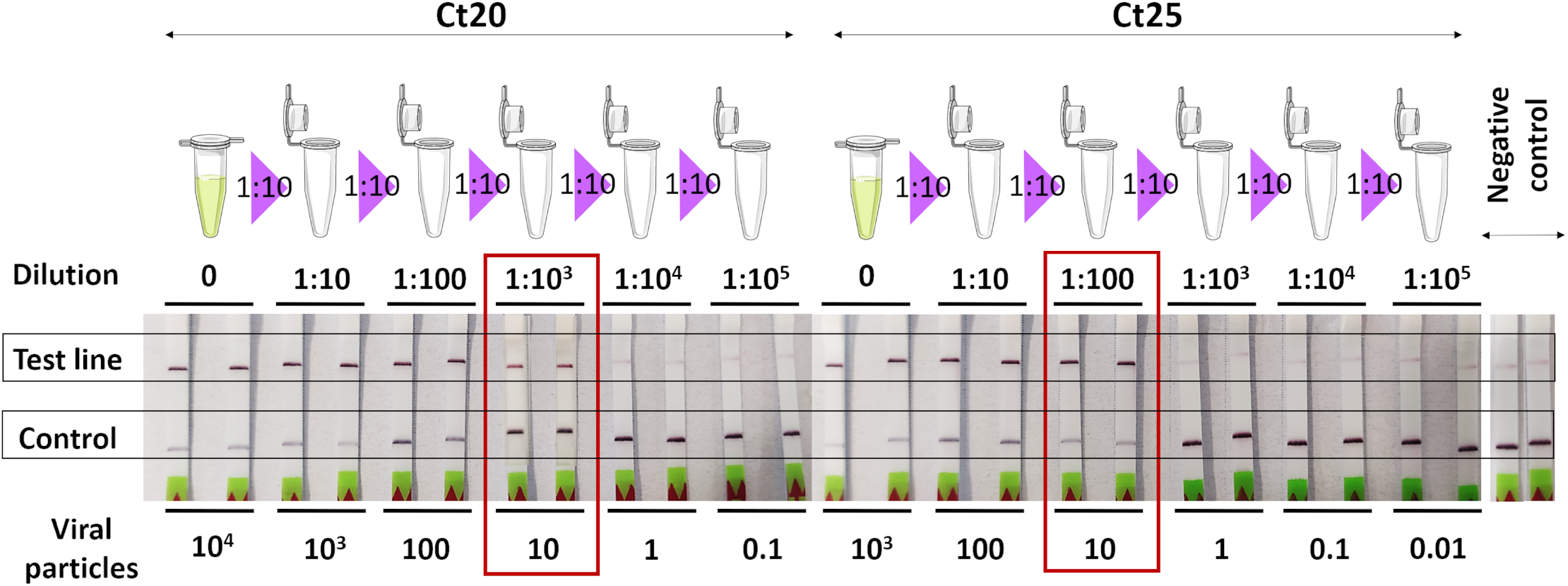
LOD assay for SARS-CoV-2 detection with N2 gene as target using serial dilutions (1:10) from two samples with different Cts (Ct20 and Ct25, respectively).

**Figure 3.**
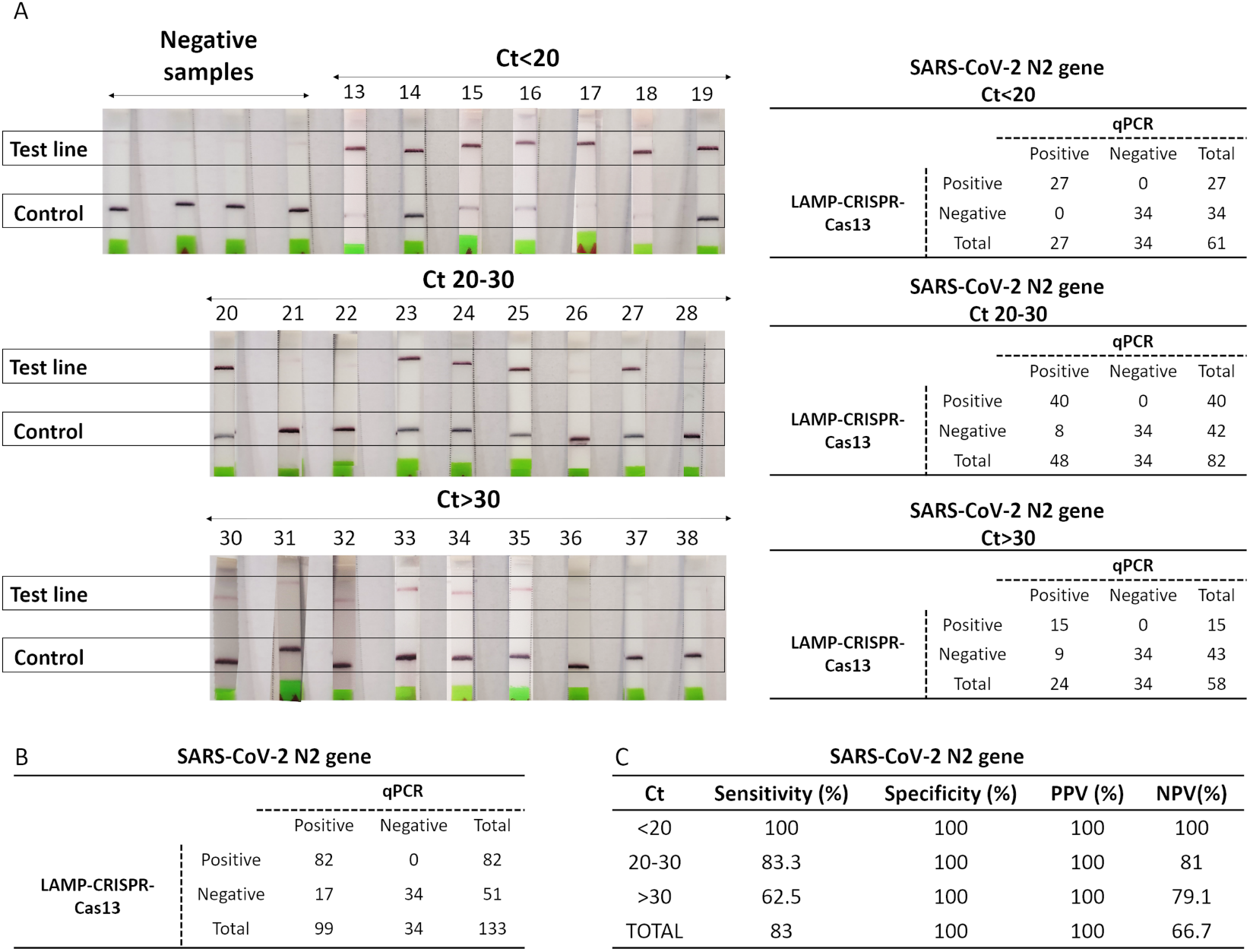
A. Strips for SARS-CoV-2 detection using samples with Cts from 13 to 38 and negative samples as negative controls with numerical results for each interval of Cts (Ct<20, Ct 20-30 and Ct>30); B. Results obtained from N2 gene SARS-CoV-2 detection; C. Table containing specificity, sensitivity, PPV and NPV LAMP-CRISPR-Cas13 technique values from figure 4B data processing.

**Figure 4.**
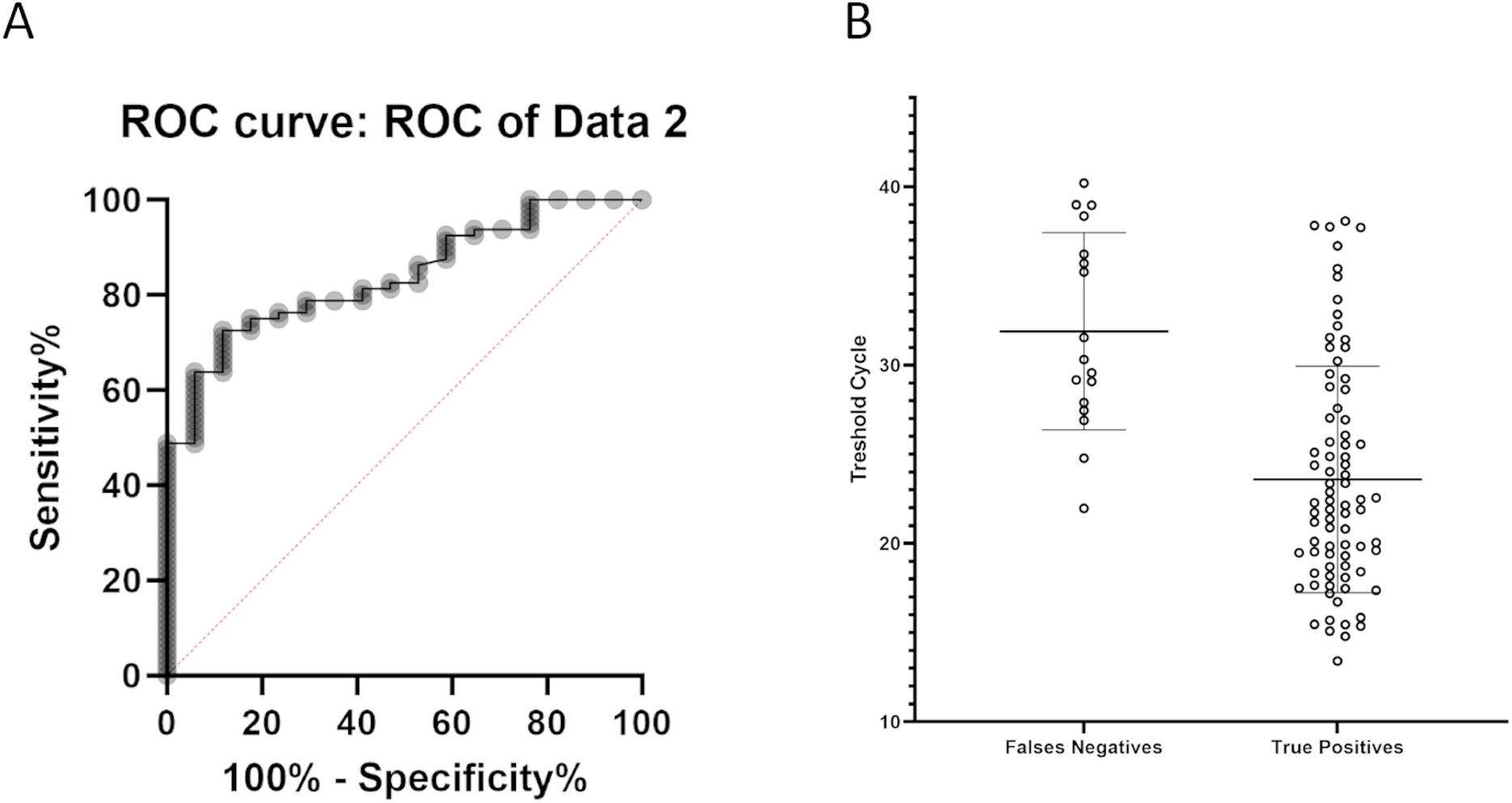
A. ROC curve for RT-LAMP-CRISPR-Cas13a technology; B. Scatter plot of two groups: false negative detections and true positive detections with RT-LAMP-CRISPR-Cas13a vs Ct of respective samples.

## Discussion

Study of the state of the art revealed that greater efforts must be made to innovate in diagnostic methods, as Bhatt A. et al. [15] found 1286 papers related to RT-LAMP and CRISPR for SARS-CoV-2 diagnosis, in contrast to the 7000 studies involving qPCR (surprisingly only 98 of these applied RT-LAMP integrated with CRISPR-Cas technology). This indicates that efforts should also be focused on developing more efficient RT-LAMP-CRISPR-Cas protocols without purification of RNA, which would reduce the cost of the testing and also produce results faster. Only 3 out of the 20 papers reviewed did not use an RNA extraction kit [16, 17, 27]. In addition, there are several advantages to the application of Cas13 endonuclease, as it has been reported to be more specific than other effector proteins [11].

In this work, our research group has developed an RT-LAMP-CRISPR-Cas13a protocol for diagnosing SARS-CoV-2 infection with an LOD of 10 viral copies, which is similar to the LOD of the qPCR method, considered the gold standard for diagnosis of COVID-19 [38]. Furthermore, it has been reported that the qPCR for SARS-CoV-2 detection has a limited sensitivity of 60-71 % [39], while the RT-LAMP-CRISPR-Cas13a technology increased this value significantly, up to 83 %.

Considering the criteria recommended by the WHO, the novel technique fulfils the three key features of accuracy, accessibility and affordability. This is because, on the one hand, it showed an accuracy ((TP+TN)/total) of 87.2 %, and, on the other hand, it is both accessible and affordable as neither specific equipment nor trained personnel are required, and the amount of enzymes needed per reaction is quite low.

Comparing our results on sensitivity, specificity, PPV and NPV with those obtained in previous studies, we found that the specificity and PPV values of the RT-LAMP-CRISPR-Cas13a technology were higher than those from 7 out of 10 RT-LAMP papers reviewed [19-25] and in one case the sensitivity of the novel technique was even higher [18]. Moreover, this technique showed higher sensitivity and NPV values than those from 2 out of 10 RT-LAMP-CRISPR papers reviewed which applied RNA extraction kit on the clinical samples [33, 35]. Among the others, 7 out of 8 studies used DNA target-endonuclease effectors, and thus higher sensitivity could be obtained due to the intrinsic stability of DNA in contrast to RNA molecules. The lower sensitivity of the RT-LAMP-CRISPR-Cas13a protocol (83 %) than described in a previous study (97.4 %) could be explained by the fact that the researchers used an RNA extraction kit (Direct-zol kit), so that the RNA was purified and concentrated, and also the results were revealed by fluorescence [26].

ROC analysis has become a popular method for evaluating the accuracy of medical diagnostic systems, as it provides accurate indices for the techniques tested that are not distorted by fluctuations caused by the use of arbitrarily chosen decision criteria or cut-off points [40]. The AUC determines the inherent ability of the test to correctly identify a person as infected or not, where an AUC value of 0.5 indicates an absence of capacity of discrimination between infected and healthy populations, a value of 0.5-0.7 is related to unsatisfactory discrimination; ability is acceptable when value is between 0.7-0.8, it is excellent for values contained in the ratio 0.8-0.9 and perfect when the AUC is close to 1 [41]. The value of the AUC ROC curve calculated by statistical analysis validated our RT-LAMP-CRISPR-Cas13a technique as a reliable diagnostic method. Furthermore, results shown in the scatter plot figure indicate that this protocol provides less accurate diagnostics when viral loads are low. However, we should bear in mind that at this stage of infection, individuals present almost no risk of being contagious [42, 43].

In summary, the high levels of specificity, sensitivity, PPV and NPV obtained with this promising protocol working with RNA-extraction kit free samples, places LAMP-CRISPR-Cas13a technology at the top of rapid and specific diagnostic methods for infectious diseases. Thus, this technique could be established as a diagnostic tool for detecting other infectious diseases, such as those caused by multiresistant pathogens. However, Cas13 detection methods should be optimized to enable direct diagnosis without prior amplification of nucleic acids.

## Data Availability

All data produced in the present work are contained in the manuscript

## Acknowledgment

This study was funded by grant PI19/00878 awarded to M. Tomás, within the State Plan for R+D+I 2013-2016 (National Plan for Scientific Research, Technological Development and Innovation 2008-2011) and co-financed by the ISCIII-Deputy General Directorate for Evaluation and Promotion of Research - European Regional Development Fund “A way of Making Europe” and Instituto de Salud Carlos III FEDER, Spanish Network for the Research in Infectious Diseases (REIPI, RD16/0016/0006 and RD16/CIII/0004/0002), Instituto de Salud Carlos III FEDER Experiments were funded by grant IN607A 2020/38 within the GAIN (Agencia Gallega de Innovación) COVID plan and by the Study Group on Mechanisms of Action and Resistance to Antimicrobials, GEMARA (SEIMC, http://www.seimc.org/) and finally, ESCMID grant (European Society of Clinical Microbiology and Infectious Diseases) awarded to L. Fernández-García. O. Pacios, L. Fernández-García and M. López were financially supported by the grants IN606A-2020/035, IN606B-2021/013 and IN606B-2018/008, respectively (GAIN, Xunta de Galicia). I. Bleriot was financially supported by pFIS pro-511gram (ISCIII, FI20/00302). Ethical approval was granted by the Galicia Drug Research Ethics Committee (CEIm-G) and the internal ethical from Institute of Research A Coruña (INIBIC) from Coruña hospital (HUAC) (2020/207).

